# Gastric modulation of food reward, olfaction and taste in obesity and bariatric surgery: an artificial intelligence assisted scoping review protocol

**DOI:** 10.1101/2024.04.29.24306451

**Authors:** Nina Ritsch, Camille Bourque, Frederic Bergeron, Julie-Anne Nazare, Anestis Dougkas, Sylvain Iceta

## Abstract

**Objective:** To understand the extent and nature of the available research on gastric modulation of food reward, olfaction, and taste in people with obesity or those who have undergone bariatric surgery.

**Introduction:** Bariatric surgery-induced weight loss is partially attributed to shifts in food preferences resulting from alterations in sensory perceptions and changes in reward system. The stomach’s innervation and mechanical function have been theorized to play a significant role in these modifications, as suggested by numerous preclinical studies. However, the extent and nature of these connections in clinical settings require further elucidation.

**Inclusion criteria:** This review will examine studies on the influence of gastric innervation and/or mechanical function on food reward, olfaction, and taste. Selected studies will include participants of all ages with obesity or bariatric surgery. Both observational studies and controlled experiments will be considered, while study protocols, opinion articles, letters to the editor, book chapters, oral communication or poster abstracts and systematic reviews will be excluded.

**Methods:** The search will be undertaken in MEDLINE, Embase, PsycINFO, CINAHL, Web of Science, Google Scholar, and gray literature. No date parameters will be set, and all languages will be considered. Citations will be uploaded into EndNote 20.0 and duplicates removed using Covidence. The remaining studies will be analyzed by 3 reviewers using a two-stage procedure with the ASReview python package. The full-text screening and the data extraction will be conducted by 2 reviewers on Covidence. An additional reviewer will be consulted in the event of disagreement. Tabulated results will be accompanied by a narrative summary.

## Introduction

Obesity is a global public health problem, partly attributed to the dysregulation of eating behavior. Among other things, it is associated with impairment in both the homeostatic and hedonic control of food intake ^1^. While bariatric surgery is recognized as the most effective treatment for obesity, it still results in weight failure (i.e., no weight loss or weight regain) in 20 to 30% of cases at 10 years ^2^. Weight loss, however, is not solely driven by a reduction in food intake due to decreased gastric volume. It is also linked to shifts in food preferences caused by alterations in sensory perceptions ^3,4^, and remodeling of reward circuit ^5^. Moreover, our research team has shown that the dietary preferences of patients with weight failure differ from those who have achieved successful post-operative outcomes^4^. These findings suggest that the variances observed in weight loss trajectories may be partially attributed to the diversity of responses of food intake control mechanisms.

Beyond its impacts on the hedonic regulation of food intake, bariatric surgery also deeply impacts the intestine-brain axis. It is hypothesized to normalize the composition of the intestinal microbiota and the secretion of digestive hormones ^6,7^, both of which are altered in people with obesity. Furthermore, these changes seem to correlate positively with post-operative weight loss ^6,7^. Similarly, increased blood levels of the gut hormones glicentin and oxyntomodulin are thought to be associated with changes in food preferences ^7^, and the digestive hormone GLP-1 has been suggested as a key factor in the perception of sweetness ^8^. Gut hormones and microbiota have been extensively investigated in the context of their significance in obesity ^9,10^ and bariatric surgeries ^11,12^.

Nonetheless, other mechanisms may be involved, such as gastric motor functions and innervations. The association between some of the gut hormones and the brain may be mediated by the vagus nerve, responsible for visceral interoception. It connects the digestive system, notably the stomach or intestine and their mechano- and chemoreceptors, to the brain via the nucleus of the tractus solitarius ^13^. This nucleus also receives afferents from the facial and glossopharyngeal nerves, responsible for taste sensitivity. Indeed, acute stimulation of the vagus nerve in depressed subjects seems to lead to an increase in the perceived intensity of sweet taste ^14^. Paradoxically, the same study suggests that this stimulation also leads to an increase in detection thresholds for sweet, sour and bitter tastes. In addition, vagus nerve would increase olfactory discrimination scores in healthy subjects ^15^, in line with the results obtained by Garcia-Diaz in 1984, who showed that acute stimulation of the vagus nerve increased electrical activity in the olfactory bulbs of rats ^16^. Also, chronic stimulation of the vagus nerve in obese guinea pigs has been shown to modify food preferences, notably by reducing consumption of sweet foods ^17^. Its chronic stimulation in obese guinea pigs induces changes in food preferences, including a reduction in the consumption of sweet foods ^17^. It also increases dopamine secretion and influences food preferences in mice ^18^. Regarding human studies, it has been shown that an acute stimulation of the vagus nerve in healthy subjects increases food reward seeking ^19^. Iatridi et al. showed in 2011 that women who liked sweet tastes had higher interoceptive capacity than those who did not ^20^. Notably, interoceptive abilities are positively associated with vagal tone ^21^. These results could partly explain the positive impact of this stimulation on the weight of obese rat ^22^ and guinea pigs ^23^. In humans, low vagal tone has been linked to obesity ^24^, while reduced interoceptive capacity has been associated with higher BMI ^25^. Figure 1 provides a summary of the connections between the vagus nerve and the stomach.

Bariatric surgery, particularly sleeve gastrectomy and Roux-en-Y gastric bypass, impact the vagus nerve by modifying its structure ^26^. Hence, it is imperative to elucidate the ramifications of this vagal restructuring on the homeostatic and hedonic mechanisms that influence food choices and their interactions. Finally, understanding their links with post-operative weight loss could promote the success of future surgery.

However, if literature is available regarding the interaction between the gut and the reward systems, it focuses on animal studies. To our knowledge, no current reviews encompass both gut innervation and motility simultaneously, nor do they consider food reward, olfaction, and taste within the same review. Furthermore, these mechanisms altogether have been insufficiently studied in the context of obesity. Therefore, conducting a scoping review in this domain would provide a comprehensive overview of current knowledge and help to identify the gaps to fill in future studies.

The primary objective of this scoping review is to systematically examine and map the existing literature on gastric modulation of food reward and preference, olfaction, and taste in people with obesity or those who have undergone bariatric surgery. Specifically, it will examine i) mechanisms related to gastric innervation encompassing both sympathetic (i.e., splanchnic nerve), and parasympathetic (i.e., vagus nerve) innervation; and ii) mechanisms related to gastric satiation, dilatation and emptying. A secondary aim of this review will be to determine whether bariatric surgery can have an impact on these mechanisms. Lastly, we will summarize the tools used in the reviewed literature for assessing reward, olfaction, and taste. Based on the findings of this scoping review, we will identify gaps in the evidence for future research and may undertake a subsequent systematic review with quantitative synthesis (e.g., investigating aspects such as the impact of vagal nerve modulation on food preferences).

## Review question

What is the extent and nature of the available research literature related to gastric modulation of food reward, olfaction and taste in people who have obesity?

- Which mechanisms related to gastric innervation impact on food reward, olfaction, and taste have been studied in human studies on obesity?
- What published literature exists regarding gastric mobility, dilatation, and emptying on food reward, olfaction, and taste.
- What level of investigation exists regarding bariatric surgery’s impact on these mechanisms?
- Which tools and outcome measures have been used to explore the gastric modulation of food reward, olfaction, and taste?

## Eligibility criteria

### Participants

Articles including participants of any age and with obesity (Body Mass Index ≥ 30 kg.m^-2^) will be included in this review as well as participants who underwent bariatric surgery (e.g., sleeve gastrectomy, Roux-en-Y Bypass or Biliopancreatic derivation).

### Concept

This review will examine studies on food reward, olfaction, and taste, whether these concepts were studied together or separately in studies.

Food intake is partly driven by food reward, which can be conceptualized as a system that assigns a hedonic value to food and generates motivation for food intake. Food reward intersects external inputs from the sensory aspects of food and internal signals from tissues, nutrients, and hormones from the gastrointestinal tract to guide eating behavior ^9^. Rather than a unitary construct, food reward consists of distinct subcomponents, which remains a subject of controversy. For the purposes of this review, we will categorize food reward in a two dimensions model: Incentive Salience or Wanting (effort-based food-directed behavior), and Hedonic Evaluation or Liking (palatability, food preference)^27^. A variety of methods have been used to measure food reward. The most common measure of food reward is food liking, food wanting (e.g., implicit, and explicit wanting) and self-reported desire to eat a specific food. Mostly self-reported tools are used such as visual analog scales (VAS). In addition, more advanced questionnaires or computerized tasks can also be used (e.g., visual-probe task, forced choice tasks, or implicit association task). Although not a direct measure of food reward, food choices and preferences are commonly used as an indicator of variation in food reward ^28^. Food preferences include the qualitative evaluation of foods, and quantitative food preference measurements (e.g., food preferences or frequency questionnaires).

Olfaction is defined at a peripheral level as “olfaction sensitivity,” where an odor is detected above a certain threshold that is specific to each individual. At a central level, olfaction gathers “indicators of olfactory performances” with odors identification abilities, and “odors perceptual ratings” with subjective characterizations of smells (intensity, familiarity, edibility). The hedonic aspect of olfactory perception is described by the perceived pleasantness of an odor.

Similarly, to olfaction, taste corresponds to “gustatory sensitivity,” where a taste is detected above a certain threshold that is specific to each individual. It also brings together “indicators of gustatory performances” with taste identification abilities, and “tastes perceptual ratings” with subjective perceptions of tastes (intensity, familiarity). The hedonic aspect of gustatory perception is described by the perceived pleasantness of taste.

### Context

This scoping review will include studies that explored the modulation or control of food reward, olfaction, and taste by the stomach. The stomach is a complex organ with mechanical and endocrine functions. The stomach’s four key digestive functions are reservoir function, acid secretion, enzyme secretion and involvement in gastrointestinal motility. The stomach’s reservoir capacity (around 1 to 2 liters) allows its volume to increase, while internal pressure increases modestly (i.e., gastric interoception). Stomach contraction and relaxation are partly mediated by a vasovagal response. Enteroendocrine hormones have pleiotropic effects in both central and peripheral systems involved in energy homeostasis and obesity regulation^9^. As many reviews have already been conducted regarding hormonal modulation and even if endocrine functions were of major interest, we decided to focus on gastric mechanical functions and gastric innervation, since we believe these components are less known.

Observational studies, as well as controlled experiments will be considered. This scoping review will include studies conducted in all countries and territories. Information about the location of each study will be integrated in the data extraction and the findings will be presented by continents or relevant geographic regions. Only studies in humans will be considered.

### Types of Sources

This scoping review will consider peer-reviewed quantitative, qualitative, and mixed methods study designs, and relevant gray literature if it meets the inclusion criteria. Observational studies including prospective and retrospective cohort studies, case-control studies and cross-sectional studies will be considered for inclusion.

The review will exclude study protocols, opinion articles, letters to the editor, book chapters, oral or poster abstracts, and systematic reviews. All sources that are included in the review will have their references examined for other potential studies to include.

## Methods

The proposed scoping review will be conducted in accordance with the JBI methodology for scoping reviews ^29^. This scoping review project’s protocol was first registered in The Open Science Framework (OSG) website on 2023/09/18. All materials, including the original protocol, revised protocol, and database search results, will be accessible through OSF registration website (https://osf.io/).

### Search strategy

The development of the three-stage research strategy will follow JBI’s recommendations. Step 1, the research team will identify a series of terms that will be used to launch an initial search in PubMed. Using data mining software (PubReMiner v1.31), an analysis of the words contained in the titles and abstracts of the articles, as well as the indexing terms, will follow.

The final search strategy will be formulated according to PCC elements: population, concepts, and context. In our research, the target population is people suffering from obesity (BMI ≥ 30 kg.m-2) and people who had undergone bariatric surgery (e.g., Obesity OR Bariatric Surgery OR Sleeve Gastrectomy OR Roux-en-Y Bypass). Our concept of interest in this review is food reward (e.g., Feeding Behavior OR Food Preferences OR Food Reward OR Liking OR Wanting OR Olfaction OR Olfactory OR Taste OR Gustatory). Finally, the last concept will focus on gastric related terms (Gastr* OR Stomach OR Gastric Dilatation OR Interoception).

In step 2, the final search strategy will be adapted and performed in the following databases: Medline (Ovid), Embase, (Embase.com), CINAHL (EBSCO), PsycInfo (Ovid) and Web of Science. In addition, Google Scholar search engine and Hal Documentation (https://hal.science/) will be searched to retrieve gray literature. No restrictions will be set to ensure no relevant sources are missed. All languages will be included to reduce the risk of missing relevant sources. Languages other than English will be translated by colleagues who are fluent in the language or through Google Translate or DeepL. A draft Medline search strategy can be found in Appendix 1.

In step 3, reference lists of all included articles will be screened to identify additional relevant articles. If key authors publishing on the review topics are identified, a specific search will be conducted on these authors.

### Study/Source of Evidence Selection

Following the search, all identified citations will be collated and uploaded into EndNote 20.0 (Clarivate Analytics, PA, USA) and duplicates removed using Covidence. The remaining studies will be analyzed using a two-stage procedure with the ASReview Lab v1.2.1 python package ^30^.

In the first stage, screening will be conducted independently on titles and abstracts by two coders (NR and CB) with an AI-assistant tool, ASReview, which adopted Natural Language Processing technique ^30^. At the beginning, abstracts will be randomly selected to train the two coders. A coder could end the screening if ASReview yielded 1% of the total number of abstract continuous irrelevant abstracts (with a minimum of 10% of total abstracts screened). The first round of screening will use default ASReview parameters (i.e., Term Frequency-Inverse Document Frequency, TF-IDF as feature extraction technique and Naive Bayes as classifier) and each reviewer will use a different set of records as prior knowledge. The consistency of the two coders’ decisions had to exceed 75% ^31^. Another round of screening for training purposes will be considered if the first consistency is not satisfied. As the best active learning criterion remains controversial, we will provide a more heuristic approach by using a ‘switching strategy’. To do so, a third reviewer (SI) will proceed to a new screening round using a more advanced and intense computational model active learning model (Doc2Vec as feature extraction technique and the fully connected neural network with 2 hidden layers as classifier). The 20 relevant and 20 irrelevant abstracts labeled in the first screening round as prior knowledge set. Same stopping criterion will be used.

The second stage focuses on the full-text screening and will be conducted on Covidence. Like the abstract screening, according to JBI Manual for Evidence Synthesis, a pilot training procedure is recommended before the formal full-text screening ^31^. Inclusive/exclusive decisions in the pilot stage of two coders will be compared until a 90% agreement rate is obtained. The final consistency between the two coders will be reported. Reasons for exclusion of sources of evidence at full text that do not meet the inclusion criteria will be recorded and reported in the scoping review. Any disagreements that arise between the reviewers at each stage of the selection process will be resolved through discussion, or with an additional reviewer (SI or JAN experts in obesity and food reward). The results of the search and the study inclusion process will be reported in full in the final scoping review and presented in a Preferred Reporting Items for Systematic Reviews and Meta-analysis extension for scoping review (PRISMA-ScR) flow diagram ^32^.

### Data Extraction

Data will be extracted from papers included in the scoping review by two independent reviewers using Covidence as a data extraction tool. The data extracted will include specific details about the participants, concept, context, study methods and key findings relevant to the review questions.

A draft extraction form is provided (see Appendix II). Two independent reviewers will pilot test the data extraction tool using 10 sources that will include a mix of original research, reviews, editorial papers, opinion papers, and gray literature. The draft data extraction tool will be modified and revised as necessary during the process of extracting data from each included evidence source. Modifications will be detailed in the scoping review. Any disagreements that arise between the reviewers will be resolved through discussion with an additional reviewer (SI). If appropriate, authors of papers will be contacted (a maximum of twice) to request missing or additional data, where required.

### Data Analysis and Presentation

The data presentation will be based on JBI scoping review guidelines. The forthcoming presentation of the scoping review’s outcomes will be structured into xx distinct sections. The first section will provide detailed reporting of the search strategy results, the selection process and the characteristics of the excluded studies, clarified with the help of a visually intuitive flow chart. The following 3 sections will present in a narrative format, as well as in synthetic tabular format, the results for olfaction, taste, and food reward in people with obesity. As results for the bariatric population are expected to be scarcer, the three dimensions (i.e., olfaction, taste, and reward) will be merged into a fourth section devoted to this population. A fifth section will then focus on a narrative presentation of the assessment tools or methods used to explore olfaction, taste, and reward. Finally, we will discuss the overall results and perspectives resulting from this work.

Moreover, comprehensive data set will be presented in a tabular format, following the extraction form presented in Annex II. Tables will be available for each dimension studied: taste, olfaction, reward, and for both obesity and bariatric surgery population. Tables will be provided in the supplementary materials to avoid overloading the article.

## Data Availability

All data produced in the present study will be available upon reasonable request to the authors once the scoping review will be published.

## Acknowledgments

n/a

## Appendices

### Appendix I: Databases search strategy (draft)

Medline (Ovid)

Date of the search: XX-XX-2023

Database limit: XX

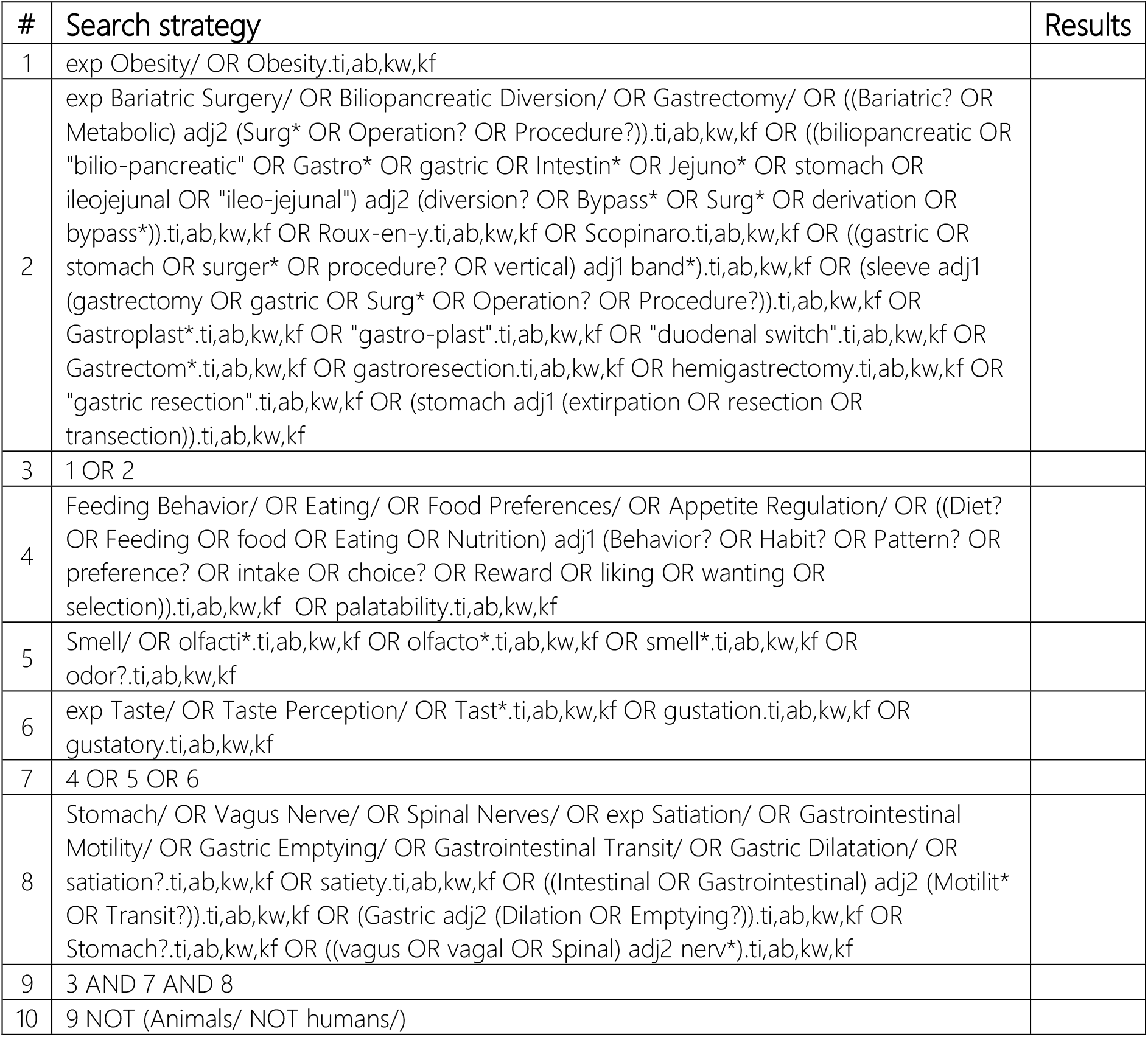

### Appendix II: Data extraction instrument

#### General information

- Author (year)
- Publication type/source (e.g., journal, thesis)
- Country
- Research design
- Study aims / objectives
- Population / Participants
- Specific inclusion / exclusion criteria

#### Population of the study

- Numbers of participants (n)
- Male – Female (%)
- Time of menstrual cycle
- Age (years)
- Type 2 Diabetes (%)
- BMI (kg/m^2^)
- Waist circumference (cm)
- Waist/Hip ratio
- Visceral fat mass (kg or %)
- Subcutaneous fat mass (kg)
- Total fat mass (kg)

*In addition, for bariatric surgery:*

- Type of surgery (sleeve gastrectomy, Roux-en-Y gastric bypass, biliopancreatic diversion)
- Time after intervention (months)
- BMI before the surgery (kg/m^2^)
- Weight loss outcome (e.g., %EWL, %TWL)
- Lowest BMI since surgery (kg/m^2^)

#### Intervention and outcomes

- Intervention type (if any)
- Outcome measure
- Methodology
- Taste: taste modality (sweet, salty, sour, bitter, umami etc.); stimuli and concentration (sucrose, quinine etc.); and gustatory function (detection, identification, intensity, pleasantness etc.)
- Olfaction: olfactory function (detection, identification, intensity, pleasantness etc.) and methods (Sniffin Sticks, ETOC (European Tests of Olfactory Capabilities) etc.)
- Reward: type of reward measure, other measure (food frequency questionnaires, 24h recall etc.)
- Key results

## References

1. Amin T, Mercer JG. Hunger and Satiety Mechanisms and Their Potential Exploitation in the Regulation of Food Intake. Curr Obes Rep. 2016; 5(1):106–12.

2. Shah M, Simha V, Garg A. Review: long-term impact of bariatric surgery on body weight, comorbidities, and nutritional status. J Clin Endocrinol Metab. 2006; 91(11):4223–31.

3. Guyot E, Dougkas A, Nazare JA, Bagot S, Disse E, Iceta S. A systematic review and meta-analyses of food preference modifications after bariatric surgery. Obes Rev. 2021; 22(10):e13315.

4. Guyot E, Dougkas A, Robert M, Nazare JA, Iceta S, Disse E. Food Preferences and Their Perceived Changes Before and After Bariatric Surgery: a Cross-sectional Study. Obes Surg. 2021; 31(7):3075–82.

5. Guyot E, Nazare JA, Oustric P, Robert M, Disse E, Dougkas A, et al. Food Reward after Bariatric Surgery and Weight Loss Outcomes: An Exploratory Study. Nutrients. 2022; 14(3).

6. Gasmi A, Bjørklund G, Mujawdiya PK, Semenova Y, Dosa A, Piscopo S, et al. Gut microbiota in bariatric surgery. Crit Rev Food Sci Nutr. 2022:1–16.

7. Nielsen MS, Ritz C, Wewer Albrechtsen NJ, Holst JJ, le Roux CW, Sjödin A. Oxyntomodulin and Glicentin May Predict the Effect of Bariatric Surgery on Food Preferences and Weight Loss. J Clin Endocrinol Metab. 2020; 105(4).

8. Jensterle M, DeVries JH, Battelino T, Battelino S, Yildiz B, Janez A. Glucagon-like peptide-1, a matter of taste? Rev Endocr Metab Disord. 2021; 22(4):763–75.

9. Crooks B, Stamataki NS, McLaughlin JT. Appetite, the enteroendocrine system, gastrointestinal disease and obesity. Proc Nutr Soc. 2021; 80(1):50–8.

10. Torres-Fuentes C, Schellekens H, Dinan TG, Cryan JF. The microbiota-gut-brain axis in obesity. Lancet Gastroenterol Hepatol. 2017; 2(10):747–56.

11. Debédat J, Clément K, Aron-Wisnewsky J. Gut Microbiota Dysbiosis in Human Obesity: Impact of Bariatric Surgery. Curr Obes Rep. 2019; 8(3):229–42.

12. Guerrero-Hreins E, Foldi CJ, Oldfield BJ, Stefanidis A, Sumithran P, Brown RM. Gut-brain mechanisms underlying changes in disordered eating behaviour after bariatric surgery: a review. Rev Endocr Metab Disord. 2022; 23(4):733–51.

13. Quadt L, Critchley HD, Garfinkel SN. The neurobiology of interoception in health and disease. Ann N Y Acad Sci. 2018; 1428(1):112–28.

14. Sperling W, Biermann T, Spannenberger R, Clepce M, Padberg F, Reulbach U, et al. Changes in gustatory perceptions of patients with major depression treated with vagus nerve stimulation (VNS). Pharmacopsychiatry. 2011; 44(2):67–71.

15. Maharjan A, Wang E, Peng M, Cakmak YO. Improvement of Olfactory Function With High Frequency Non-invasive Auricular Electrostimulation in Healthy Humans. Front Neurosci. 2018; 12:225.

16. García-Díaz DE, Aguilar-Baturoni HU, Guevara-Aguilar R, Wayner MJ. Vagus nerve stimulation modifies the electrical activity of the olfactory bulb. Brain Res Bull. 1984; 12(5):529–37.

17. Val-Laillet D, Biraben A, Randuineau G, Malbert CH. Chronic vagus nerve stimulation decreased weight gain, food consumption and sweet craving in adult obese minipigs. Appetite. 2010; 55(2):245–52.

18. Han W, Tellez LA, Perkins MH, Perez IO, Qu T, Ferreira J, et al. A Neural Circuit for Gut-Induced Reward. Cell. 2018; 175(3):665-78.e23.

19. Neuser MP, Teckentrup V, Kühnel A, Hallschmid M, Walter M, Kroemer NB. Vagus nerve stimulation boosts the drive to work for rewards. Nat Commun. 2020; 11(1):3555.

20. Iatridi V, Quadt L, Hayes JE, Garfinkel SN, Yeomans MR. Female sweet-likers have enhanced cross-modal interoceptive abilities. Appetite. 2021; 165:105290.

21. Richter F, García AM, Rodriguez Arriagada N, Yoris A, Birba A, Huepe D, et al. Behavioral and neurophysiological signatures of interoceptive enhancements following vagus nerve stimulation. Hum Brain Mapp. 2021; 42(5):1227–42.

22. Yao G, Kang L, Li J, Long Y, Wei H, Ferreira CA, et al. Effective weight control via an implanted self-powered vagus nerve stimulation device. Nat Commun. 2018; 9(1):5349.

23. Sobocki J, Fourtanier G, Estany J, Otal P. Does vagal nerve stimulation affect body composition and metabolism? Experimental study of a new potential technique in bariatric surgery. Surgery. 2006; 139(2):209–16.

24. Riva P, Martini G, Rabbia F, Milan A, Paglieri C, Chiandussi L, et al. Obesity and autonomic function in adolescence. Clin Exp Hypertens. 2001; 23(1-2):57–67.

25. Robinson E, Foote G, Smith J, Higgs S, Jones A. Interoception and obesity: a systematic review and meta-analysis of the relationship between interoception and BMI. Int J Obes (Lond). 2021; 45(12):2515–26.

26. Ballsmider LA, Vaughn AC, David M, Hajnal A, Di Lorenzo PM, Czaja K. Sleeve gastrectomy and Roux-en-Y gastric bypass alter the gut-brain communication. Neural Plast. 2015; 2015:601985.

27. Berridge KC, Robinson TE, Aldridge JW. Dissecting components of reward: ‘liking’, ‘wanting’, and learning. Curr Opin Pharmacol. 2009; 9(1):65–73.

28. Recio-Román A, Recio-Menéndez M, Román-González MV. Food Reward and Food Choice. An Inquiry Through The Liking and Wanting Model. Nutrients. 2020; 12(3).

29. Peters MD, Marnie C, Tricco AC, Pollock D, Munn Z, Alexander L, et al. Updated methodological guidance for the conduct of scoping reviews. JBI evidence synthesis. 2020; 18(10):2119–26.

30. Van De Schoot R, De Bruin J, Schram R, Zahedi P, De Boer J, Weijdema F, et al. An open source machine learning framework for efficient and transparent systematic reviews. Nature machine intelligence. 2021; 3(2):125–33.

31. Aromataris E, Munn Z. JBI manual for evidence synthesis. Joanna Briggs Institute; 2020.

32. Tricco AC, Lillie E, Zarin W, O’Brien KK, Colquhoun H, Levac D, et al. PRISMA Extension for Scoping Reviews (PRISMA-ScR): Checklist and Explanation. Ann Intern Med. 2018; 169(7):467–73.

